# Role of Intravenous Tranexamic Acid in Immediate Breast Reconstruction with an Extended Latissimus Dorsi Flap: A Retrospective Cohort Study

**DOI:** 10.64898/2026.11.14.26368090

**Authors:** Zhuoyue Li, Jian Ni, Shuqin Wang

**Author notes:** Corresponding Author (Ni Jian); (Shuqin wang).

## Abstract

**Background:** The purpose of this study was to assess the impact of intravenous tranexamic acid (TXA) on postoperative drainage, laboratory parameters, and complications in patients undergoing immediate extended latissimus dorsi (ELD) flap breast reconstruction.

**Methods:** This retrospective cohort study included 82 patients who underwent immediate ELD flap breast reconstruction between January 2019 and May 2026. The TXA group (n = 42) received 1 g of intravenous TXA at the start of surgery after its routine adoption in July 2021; the control group (n = 40) underwent surgery before this date without TXA. Drainage, perioperative hemoglobin, hematocrit, and CRP levels, and postoperative complications were assessed using multivariable linear regression, with Cox regression as a complementary time-to-event analysis.

**Results:** After adjustment for potential confounders, TXA use was associated with a shorter drain duration (β = −1.85 days, 95% CI −3.09 to −0.61; p = 0.004) and lower total drainage volume (β = −131.5 mL, 95% CI −247.2 to −15.8; p = 0.027). The adjusted Cox model also showed previous drain removal in the TXA group (HR = 2.59, 95% CI: 1.50-4.46; p < 0.001). Hematoma was found in 0 patients in the TXA group and 6 patients in the control group (p = 0.011); this result is to be regarded as descriptive and hypothesis-generating.

**Conclusion:** This study provides some evidence that intravenous TXA reduces postoperative drainage and hematoma rates after immediate ELD flap breast reconstruction; however, prospective randomized studies are necessary to validate its safety profile and define its exact role.

## 1. Introduction

Breast reconstruction is now part and parcel of the overall treatment of breast cancer, and it plays a significant role in patient satisfaction with body image as well as the objectives of multidisciplinary oncological care^1^. Recent research has emphasized the use of the extended latissimus dorsi (ELD) flap as an effective donor site due to its high tissue volume, predictable vascularity, and strong blood supply^2^. Several factors have led to renewed interest in the ELD flap, such as the possibility of high-volume fat grafting to increase the volume of the flap, quilting sutures to minimize the occurrence of seroma formation after surgery, and modifications in reimbursement policies regarding free tissue transfer^3^.

Asian women tend to possess smaller body habitus and lower body mass index (BMI), which can restrict the choice of appropriate donor sites and pose a challenge in choosing reconstructive options to smaller breasts^4^. Moreover, other patients might not want to use implantable foreign materials due to the fear of the complications associated with implants, and the ELD flap can be a good choice in autologous breast reconstruction^5^. Nevertheless, this method is still linked to a number of postoperative complications, such as donor-site seroma, extended drainage, and wound infection^6,7^. These complications can be caused by the large amount of tissue dissection during flap harvesting, the disturbance of blood and lymphatic vessels, and insufficient intraoperative hemostasis. As a result, prevention and treatment of hematoma and seroma at the ELD flap donor site is still a significant issue to plastic surgeons^8^.

Tranexamic acid (TXA) is an antifibrinolytic agent that is extensively applied in different surgical specialties due to its proven safety and effectiveness in preventing intraoperative blood loss and postoperative bleeding^9,10^. It has also become a more popular procedure in breast plastic surgery, such as reduction mammoplasty and breast reconstruction using implants. Interestingly, Weissler et al. found a much lower incidence of postoperative hematoma in patients who received TXA compared to the control group after undergoing breast reconstruction using implants^11^.

Despite the fact that a number of studies have shown that TXA can help decrease the occurrence of postoperative hematoma and seroma in breast reconstruction, there is limited evidence on its effectiveness and safety in ELD flap procedures. Thus, the purpose of the comparative study was to assess the perioperative effects of administration of systemic TXA in patients undergoing immediate breast reconstruction with an ELD flap, especially the postoperative drainage, surgical complications, and hematological parameters, including hemoglobin (Hb) and hematocrit levels (Hct). In particular, we compared the standard practice of use of intravenous TXA during ELD flap reconstruction to identify its possible advantages in terms of postoperative morbidity reduction and enhanced perioperative patient safety.

## 2. Materials and Methods

### 2.1 Study Design and Ethics

After obtaining approval from the Ethics Committee of Nanjing Drum Tower Hospital (2026-1069-01), we conducted a retrospective study in accordance with the Strengthening the Reporting of Observational Studies in Epidemiology (STROBE) guidelines. The sample size of the study was 82 patients who had immediate breast reconstruction with an ELD flap in one institution between January 2019 and May 2026.

### 2.2 Patient Selection and TXA Administration

Inclusion criteria were as follows: female patients aged 18-70 years who had immediate ELD flap breast reconstruction and a minimum of 1-month postoperative follow-up. The exclusion criteria were: (1) a history of coagulopathy, thromboembolic events, or hypersensitivity to TXA; and (2) inflammatory breast cancer or distant metastasis. Patients were divided into two groups according to the perioperative administration of TXA. The TXA group included patients who had surgery following the routine use of TXA in July 2021 (n = 42) and were given 1 g of intravenous TXA at the start of surgery. The control group included patients who had surgery prior to July 2021 (n = 40) and were not given TXA.

### 2.3 Surgical Technique and Postoperative Care

The same group of skilled plastic surgeons carried out all reconstructive procedures with the use of a pedicled ELD flap after mastectomy. The incision of the back skin was planned in the dorsal line of the bra strap to be easily concealed and was usually 6-8 cm wide and 20-25 cm long. The latissimus dorsi muscle was dissected only with the use of electrocautery, without local adrenaline infiltration at the site of incision. The dissection continued to Scarpa fascia, including the scapular fat covering the trapezius muscle and fat above the iliac crest, but sparing the deep fascia covering the underlying muscles to allow lymphatic drainage. The dissection was carried down to the iliac crest and up to the teres major muscle, taking great care not to damage the thoracodorsal vessels and nerve. The flap harvested was then turned through a subcutaneous tunnel to the chest. Quilting sutures were used to seal dead space at the donor site. The wound was sutured in layers, approximating the skin flaps to the deep tissue with 4-0 Vicryl (polyglactin 910) as the deep layer and 5-0 PDS (polydioxanone) as the superficial layer, with the intention of reducing the formation of seroma.

A single closed-suction drain (10 Ch, 3.3 mm) was inserted at the donor site at the end of each procedure and attached to a high-vacuum wound drainage system (600 mL Kit CH10; Medinorm Medizintechnik GmbH, Neunkirchen, Germany) with a negative pressure of 20 kPa. Nursing staff regularly documented drain output daily at 07:00 to 08:00 as a routine postoperative procedure. Continuous negative-pressure suction was kept on the drains until the daily output decreased to less than 30 mL on 3 consecutive days. Subcutaneous enoxaparin (0.4 mL, which corresponds to 4,000 IU anti-Xa activity, once a day) was used as pharmacological thromboprophylaxis on postoperative day 1 and continued until the end of the hospital stay.

### 2.4 Data Collection and Outcome Definitions

An independent researcher who was not part of the clinical decision-making process in the administration of TXA retrospectively extracted demographic, clinical, and surgical data. The main results were the duration of donor-site drainage (days) and the total amount of drain (mL). Secondary outcomes were the first 24-hour drain volume, daily drainage volumes during the first 6 postoperative days, and changes in laboratory parameters, such as Hb, Hct, and C-reactive protein (CRP), measured preoperatively and on postoperative day 1.

Postoperative complications that happened within 30 days of surgery were documented systematically and categorized based on standardized criteria:

■ **Hematoma:** Localized collection of blood at the surgical site that needs to be removed, such as bedside aspiration or surgical evacuation.
■ **Seroma:** A pool of clear fluid at the surgical site that needs to be removed, such as ultrasound-guided aspiration or evacuation in the operating room.
■ **Surgical site infection (SSI):** A surgical-site infection meeting the Centers for Disease Control and Prevention (CDC) criteria and requiring systemic antibiotic therapy.
■ **Flap necrosis:** Loss of part or all of the flaps necessitating surgical debridement.
■ **Thromboembolism:** Symptomatic deep vein thrombosis (DVT) with Doppler ultrasonography.

### 2.5 Statistical Analysis

Continuous variables are reported as mean ± standard deviation (SD) and compared with independent-samples t-tests or Welch’s t-tests, as needed. The non-parametric Mann–Whitney U test was used as a sensitivity analysis for skewed drainage data. The Fisher’s exact test was used to compare categorical variables, such as 30-day complication rates, and exact 95% confidence intervals (CIs) and conditional odds ratios (ORs) were reported where necessary.

Multivariable linear regression models that accounted for the use of TXA, age, BMI, type of reconstruction, axillary procedure, and neoadjuvant chemotherapy were prespecified as the main analyses in terms of donor-site drainage duration and total drainage volume. Sensitivity analyses were conducted using unadjusted t-tests and Wilcoxon rank-sum tests to determine how well the results were robust to distributional assumptions. Moreover, a Cox proportional hazards model was employed as a complementary analysis to describe the relationship between TXA use and time to drain removal in a time-to-event analysis. Multivariate Cox proportional hazards regression was used to assess time to drain removal, with the Efron method applied to deal with tied event times, and Kaplan-Meier curves were compared using the log-rank test.

Statistical tests were all two-sided, and p < 0.05 was regarded as statistically significant. All the analyses were conducted with the help of IBM SPSS Statistics, Version 25.0 (IBM Corp., Armonk, NY, USA). Since the omnibus F-test of the drain-duration regression model was marginally statistically significant (p = 0.056), post hoc diagnostic tests, such as the variance inflation factors and the comparison of the nested models, were conducted to determine the stability of the model. The findings of these diagnostic analyses are given in the footnote to Table 2.

## RESULTS

### 3.1 Patient Characteristics

This study included 82 patients, all of whom successfully underwent single-stage breast reconstruction using an ELD flap following nipple-sparing or skin-sparing mastectomy. Among them, 42 patients were given intravenous TXA, and 40 patients were in the control group. The baseline demographic and clinical characteristics of the two groups are summarized in Table 1.

**Table 1.** Baseline demographic and clinical characteristics.

| **Characteristic** | **TXA group (n = 42)** | **Control group (n = 40)** | ***p* value** |
|---|---|---|---|
| Age, years | 39.57±8.98 | 41.93±10.35 | 0.27 |
| BMI, kg/m<sup>2</sup> | 22.83±3.60 | 22.49±2.23 | 0.61 |
| Comorbidities |  |  |  |
| HTN | 0 (0) | 2 (5.0) | 0.24 |
| DM | 2 (4.8) | 0 (0) | 0.49 |
| Neoadjuvant chemotherapy | 21 (50.0) | 27 (67.5) | 0.12 |
| ALND | 14 (33.3) | 10 (25.0) | 0.47 |
| SLNB | 28 (66.7) | 30 (75.0) | 0.47 |
| Breast reconstruction |  |  | 0.11 |
| ELD | 31 (73.8) | 22 (55.0) |  |
| ELD+prosthesis | 11 (26.2) | 18 (45.0) |  |

Data are presented as mean ± standard deviation or n (%), as appropriate. TXA, tranexamic acid; BMI, body mass index; HTN, hypertension; DM, diabetes mellitus; ALND: Axillary lymph node dissection; SLNB: Sentinel lymph node biopsy

**Table 2.**
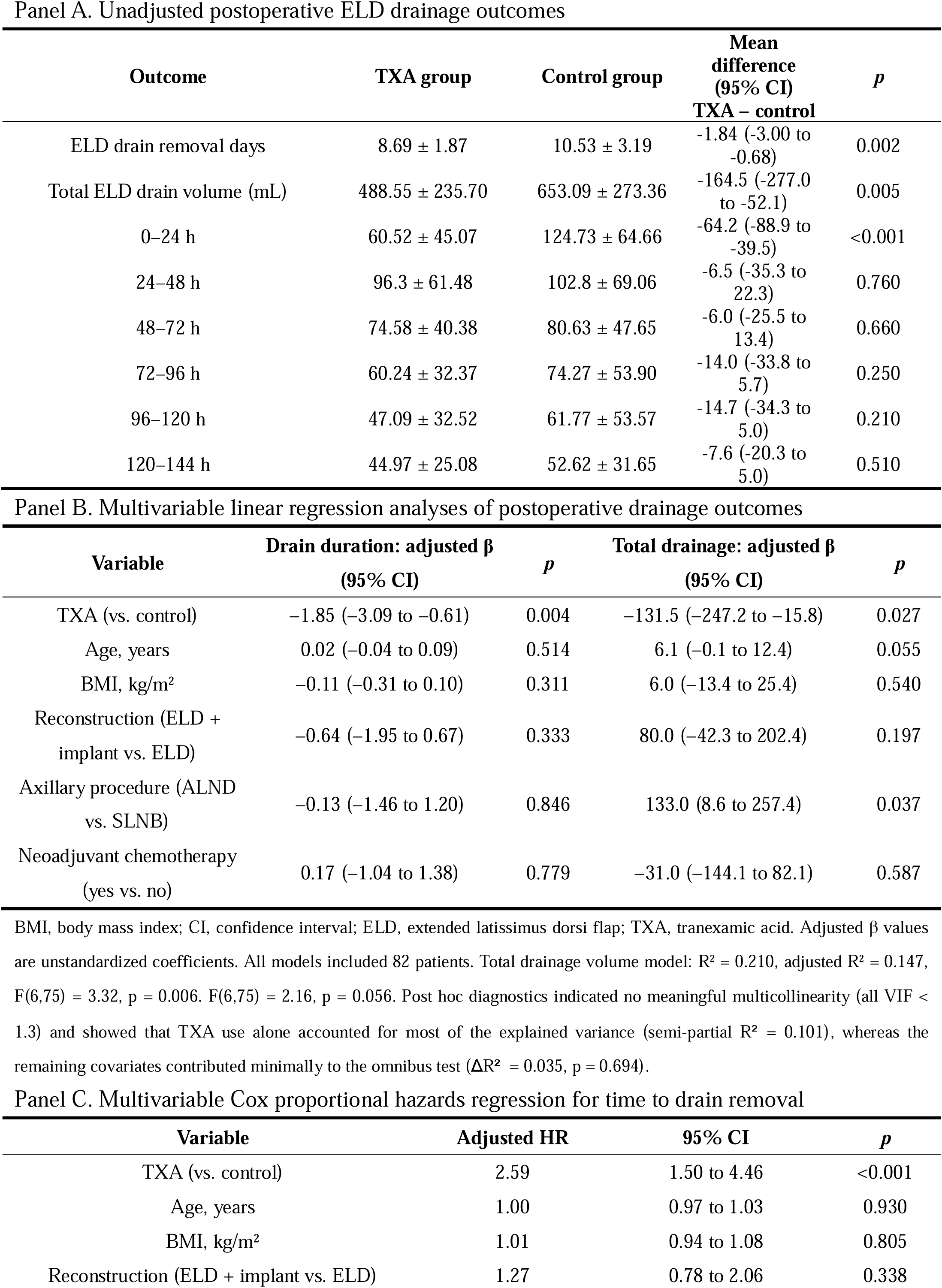
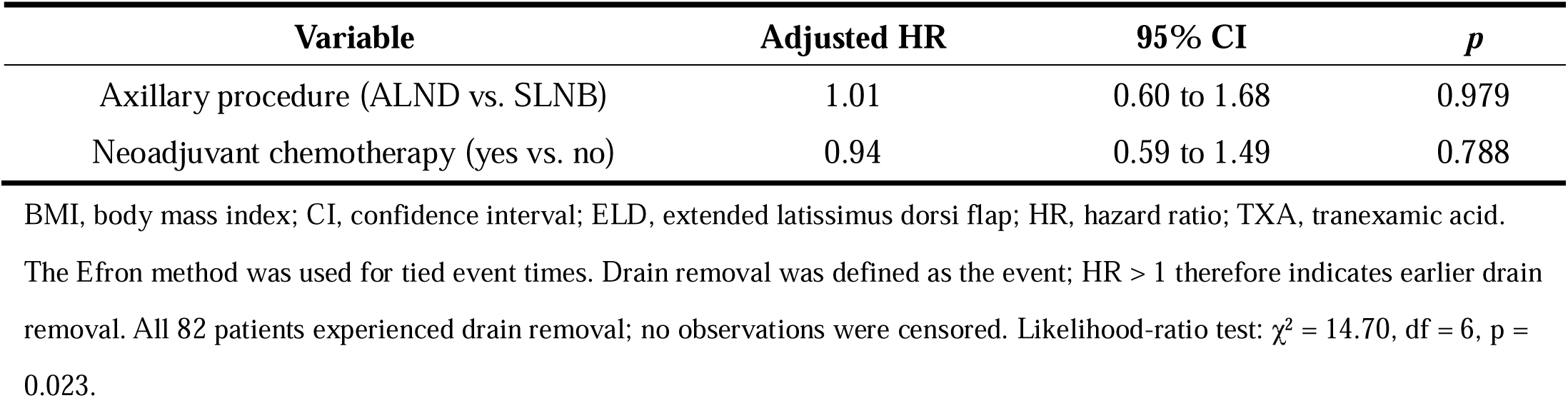
Postoperative drainage outcomes and adjusted analyses.

| Panel A. Unadjusted postoperative ELD drainage outcomes |  |  |  |  |
| **Outcome** | **TXA group** | **Control group** | **Mean difference (95% CI)<br>TXA – control** | ***p*** |
|---|---|---|---|---|
| ELD drain removal days | 8.69 ± 1.87 | 10.53 ± 3.19 | -1.84 (-3.00 to -0.68) | 0.002 |
| Total ELD drain volume (mL) | 488.55 ± 235.70 | 653.09 ± 273.36 | -164.5 (-277.0 to -52.1) | 0.005 |
| 0–24 h | 60.52 ± 45.07 | 124.73 ± 64.66 | -64.2 (-88.9 to -39.5) | <0.001 |
| 24–48 h | 96.3 ± 61.48 | 102.8 ± 69.06 | -6.5 (-35.3 to 22.3) | 0.760 |
| 48–72 h | 74.58 ± 40.38 | 80.63 ± 47.65 | -6.0 (-25.5 to 13.4) | 0.660 |
| 72–96 h | 60.24 ± 32.37 | 74.27 ± 53.90 | -14.0 (-33.8 to 5.7) | 0.250 |
| 96–120 h | 47.09 ± 32.52 | 61.77 ± 53.57 | -14.7 (-34.3 to 5.0) | 0.210 |
| 120–144 h | 44.97 ± 25.08 | 52.62 ± 31.65 | -7.6 (-20.3 to 5.0) | 0.510 |
| Panel B. Multivariable linear regression analyses of postoperative drainage outcomes |  |  |  |  |
| **Variable** | **Drain duration: adjusted β (95% CI)** | ***p*** | **Total drainage: adjusted β (95% CI)** | ***p*** |
| TXA (vs. control) | –1.85 (–3.09 to –0.61) | 0.004 | –131.5 (–247.2 to –15.8) | 0.027 |
| Age, years | 0.02 (–0.04 to 0.09) | 0.514 | 6.1 (–0.1 to 12.4) | 0.055 |
| BMI, kg/m<sup>2</sup> | –0.11 (–0.31 to 0.10) | 0.311 | 6.0 (–13.4 to 25.4) | 0.540 |
| Reconstruction (ELD + implant vs. ELD) | –0.64 (–1.95 to 0.67) | 0.333 | 80.0 (–42.3 to 202.4) | 0.197 |
| Axillary procedure (ALND vs. SLNB) | –0.13 (–1.46 to 1.20) | 0.846 | 133.0 (8.6 to 257.4) | 0.037 |
| Neoadjuvant chemotherapy (yes vs. no) | 0.17 (–1.04 to 1.38) | 0.779 | –31.0 (–144.1 to 82.1) | 0.587 |
| BMI, body mass index; CI, confidence interval; ELD, extended latissimus dorsi flap; TXA, tranexamic acid. Adjusted β values are unstandardized coefficients. All models included 82 patients. Total drainage volume model: R<sup>2</sup> = 0.210, adjusted R<sup>2</sup> = 0.147, F(6,75) = 3.32, p = 0.006. F(6,75) = 2.16, p = 0.056. Post hoc diagnostics indicated no meaningful multicollinearity (all VIF < 1.3) and showed that TXA use alone accounted for most of the explained variance (semi-partial R<sup>2</sup> = 0.101), whereas the remaining covariates contributed minimally to the omnibus test (ΔR<sup>2</sup> = 0.035, p = 0.694). |  |  |  |  |
| Panel C. Multivariable Cox proportional hazards regression for time to drain removal |  |  |  |  |
| **Variable** | **Adjusted HR** | **95% CI** | ***p*** |  |
| TXA (vs. control) | 2.59 | 1.50 to 4.46 | <0.001 |  |
| Age, years | 1.00 | 0.97 to 1.03 | 0.930 |  |
| BMI, kg/m<sup>2</sup> | 1.01 | 0.94 to 1.08 | 0.805 |  |
| Reconstruction (ELD + implant vs. ELD) | 1.27 | 0.78 to 2.06 | 0.338 |  |
| **Variable** | **Adjusted HR** | **95% CI** | ***p*** |  |
| Axillary procedure (ALND vs. SLNB) | 1.01 | 0.60 to 1.68 | 0.979 |  |
| Neoadjuvant chemotherapy (yes vs. no) | 0.94 | 0.59 to 1.49 | 0.788 |  |

BMI, body mass index; CI, confidence interval; ELD, extended latissimus dorsi flap; HR, hazard ratio; TXA, tranexamic acid. The Efron method was used for tied event times. Drain removal was defined as the event; HR > 1 therefore indicates earlier drain removal. All 82 patients experienced drain removal; no observations were censored. Likelihood-ratio test: χ<sup>2</sup> = 14.70, df = 6, p = 0.023.

### 3.2 Drainage Outcomes

TXA use was significantly related to a reduced drain time (adjusted unstandardized β = -1.85 days, 95% CI: -3.09 to -0.61; p = 0.004) and a reduced total drainage volume (adjusted unstandardized β = -131.5 mL, 95% CI: -247.2 to -15.8; p = 0.027) after age, BMI, reconstruction type, axillary procedure, and neoadjuvant chemotherapy adjustment (Table 2, Panel B). These results were in line with the unadjusted analyses. The mean donor-site drain duration was significantly shorter in the TXA group than in the control group (8.69 ± 1.87 vs. 10.53 ± 3.19 days; p = 0.002) (Figure 1A), and the total postoperative drain output was also significantly lower (488.55 ± 235.70 vs. 653.09 ± 273.36 mL; p = 0.005) (Figure 1B). The non-parametric sensitivity analyses provided the same results in terms of the drain duration (Mann-Whitney U test, p = 0.010) and the total drainage volume (Mann-Whitney U test, p = 0.014), which also supports the strength of the main adjusted estimates.

**Figure 1.**
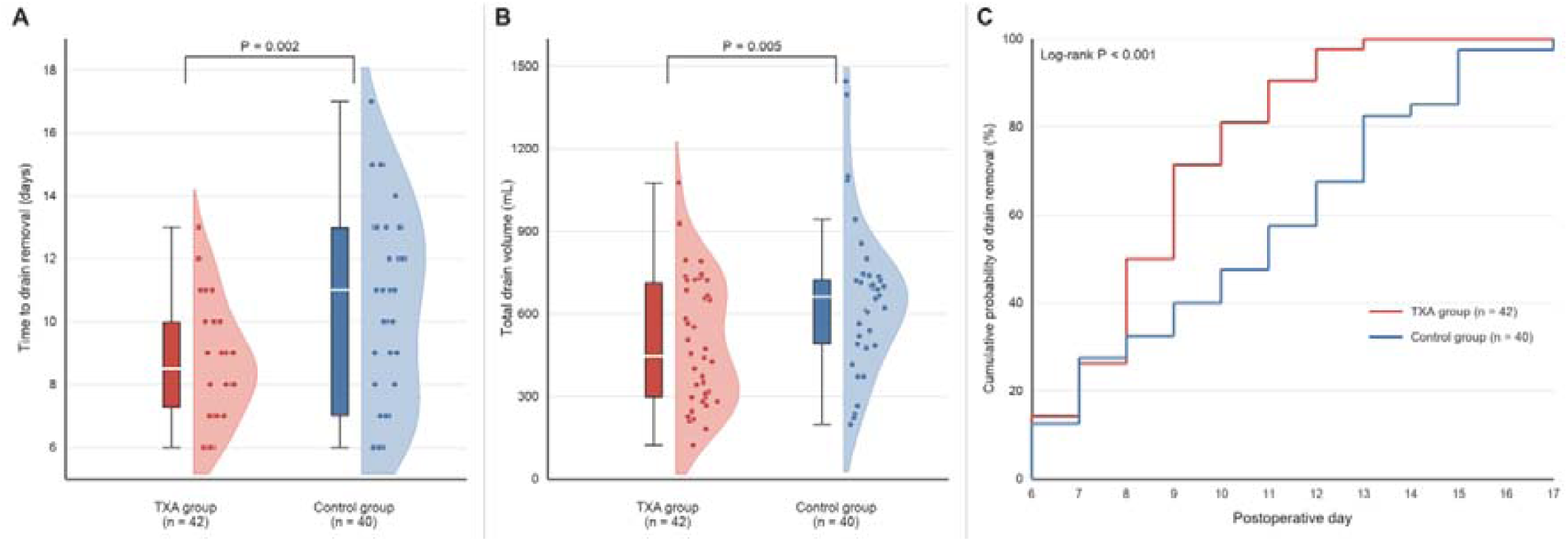
Postoperative drainage outcomes according to TXA status. (A) Time to drain removal. (B) Total postoperative drain volume. Boxes represent the interquartile range, horizontal lines indicate medians, whiskers extend to 1.5 times the interquartile range, and dots represent individual patients. (C) Kaplan–Meier cumulative drain-removal curves. HR, hazard ratio; TXA, tranexamic acid.

The greatest difference in the amount of drainage between groups was found in the first 24 hours after the operation (60.52 ± 45.07 vs. 124.73 ± 64.66 mL; p < 0.001). On the next 5 days, the mean daily drain output was numerically lower in the TXA group, but none of the between-group differences were statistically significant (Table 2, Panel A; Figure 2). On postoperative day 10, drains had been removed in 81.0% of patients in the TXA group versus 47.5% of patients in the control group (p = 0.002) (Figure 1C).

**Figure 2.**
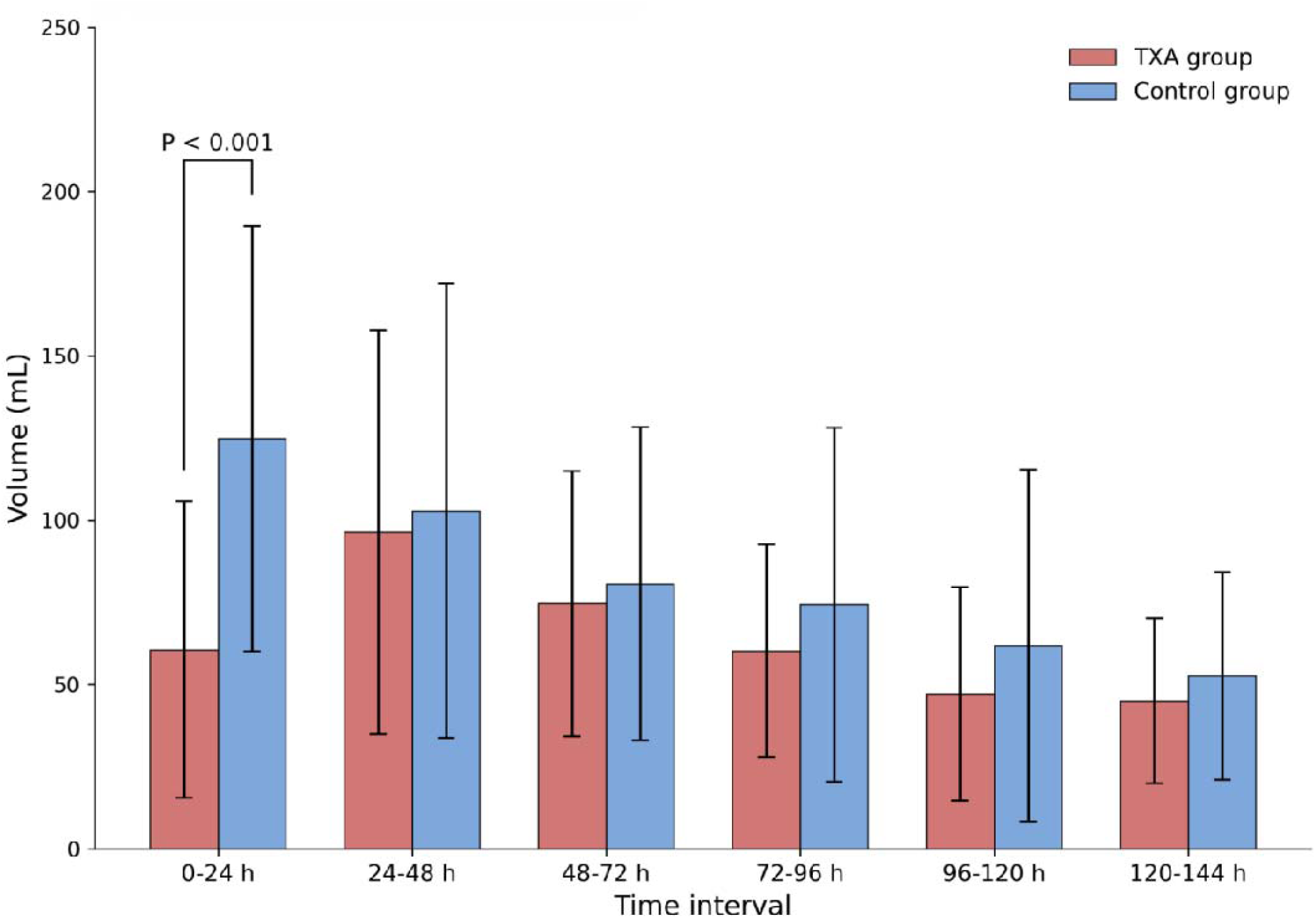
Mean daily drain volume over the first 6 postoperative days compared between the TXA group and the control group.

The adjusted Cox proportional hazards model, as a complementary time-to-event analysis, showed that the use of TXA was significantly related to an increased rate of drain removal (adjusted HR = 2.59, 95% CI: 1.50-4.46; p < 0.001), which means that patients in the TXA group had a higher rate of drain removal compared to patients in the control group (Table 2, Panel C).

### 3.3 Laboratory Findings

The research showed that the postoperative decrease in Hct levels was significantly lower in the TXA group compared to the control group (2.82% ± 2.1% vs. 4.0% ± 3.0%; p = 0.044). Likewise, the average reduction in hemoglobin (Hb) was much less in the TXA group (9.88 ± 9.63 g/L vs. 23.26 ± 12.83 g/L; p < 0.001), indicating that perioperative blood loss was less in patients who received TXA.

In terms of inflammatory markers, the preoperative CRP levels were similar in the two groups. Nevertheless, the CRP levels in the postoperative period were much lower in the TXA group compared to the control group (13.34 ± 5.01 mg/L vs. 21.26 ± 5.93 mg/L; p < 0.001). This observation could be due to decreased bleeding and/or tissue damage, and not a direct anti-inflammatory action of TXA (Table 3).

**Table 3.**
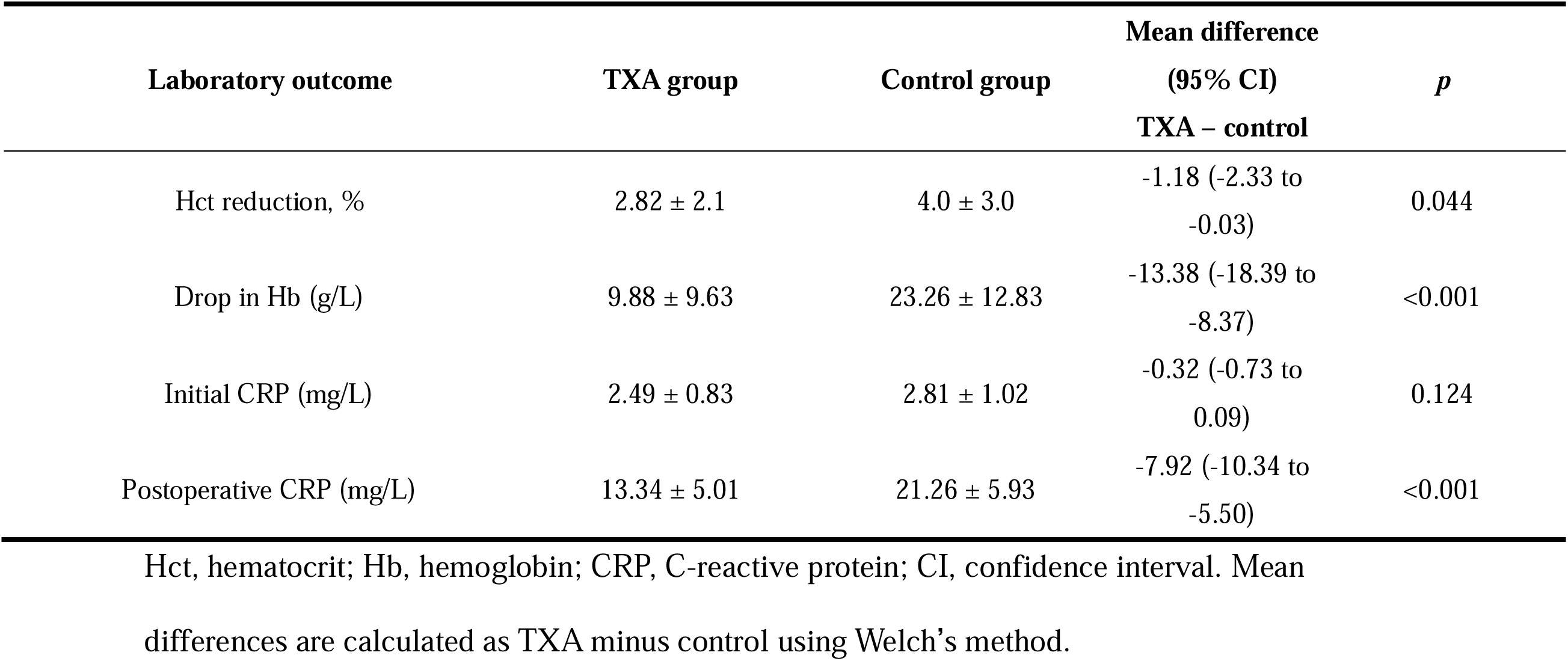
Laboratory findings.

**Table 4.**
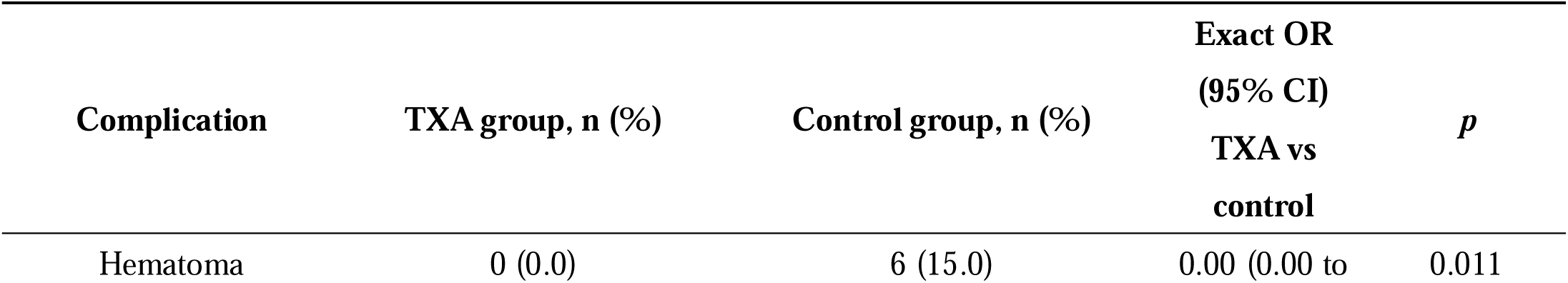
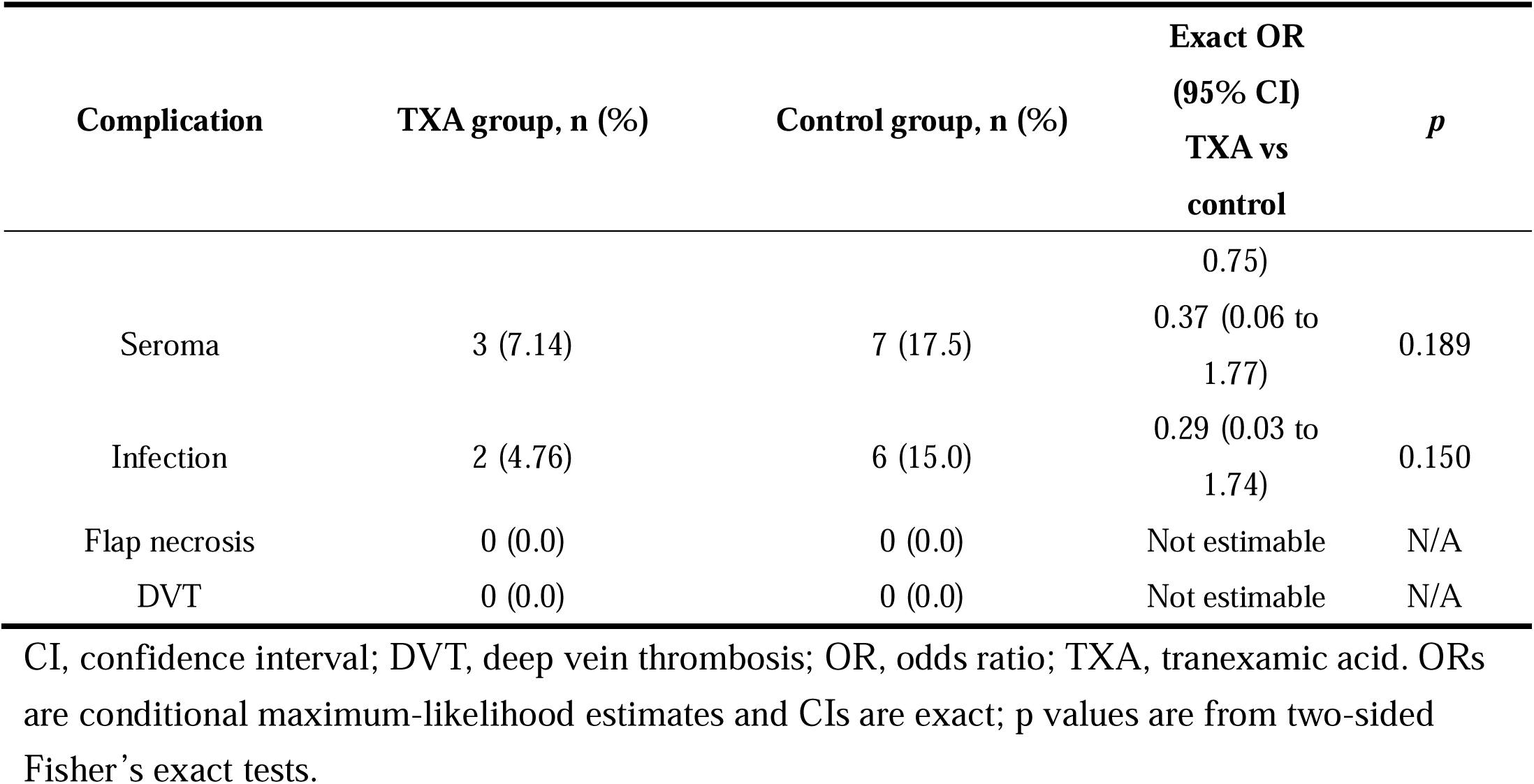
Surgical complications.

| Complication | TXA group, n (%) | Control group, n (%) | Exact OR (95% CI) TXA vs control | *p* |
|---|---|---|---|---|
| Hematoma | 0 (0.0) | 6 (15.0) | 0.00 (0.00 to 0.75) | 0.011 |
| Seroma | 3 (7.14) | 7 (17.5) | 0.37 (0.06 to 1.77) | 0.189 |
| Infection | 2 (4.76) | 6 (15.0) | 0.29 (0.03 to 1.74) | 0.150 |
| Flap necrosis | 0 (0.0) | 0 (0.0) | Not estimable | N/A |
| DVT | 0 (0.0) | 0 (0.0) | Not estimable | N/A |

CI, confidence interval; DVT, deep vein thrombosis; OR, odds ratio; TXA, tranexamic acid. ORs are conditional maximum-likelihood estimates and CIs are exact; p values are from two-sided Fisher’s exact tests.

### 3.4 Postoperative Complications

There were no significant postoperative complications such as wound dehiscence, flap necrosis, or DVT in both groups, and the overall ELD flap survival rate was 100%. Seroma was found in 3 patients (7.14%) in the TXA group and 7 patients (17.5%) in the control group, but this was not significantly different (Fisher’s exact test, p = 0.189). None of the patients in the TXA group had a hematoma, versus 6 patients (15.0%) in the control group, which is statistically significant (Fisher’s exact test, p = 0.011). The control group had all hematomas that needed intervention, with 5 patients needing bedside aspiration and 1 patient needing surgical evacuation. In the TXA group, 2 patients (4.76%) and in the control group, 6 patients (15.0%) had surgical-site infection, but the difference was not significant (Fisher’s exact test, p = 0.150). Antibiotic therapy was effective in all infections.

## Discussion

Although TXA has been widely used in different types of surgeries, the evidence on its application in breast surgery is relatively new. Most of the studies were published after 2018, although the first reports of the use of TXA in breast surgery can be traced back to 1994^12^. TXA has been used topically and systemically in breast surgery and has been linked to decreases in the rate of postoperative hematoma and seroma, and the total amount of drainage. Nevertheless, to the best of our knowledge, the existing literature does not provide any evidence on the application of a standardized systemic TXA protocol in patients who underwent mastectomy and immediate ELD flap breast reconstruction.

Donor-site complications are a significant issue after ELD flap reconstruction, especially extended drainage and postoperative fluid retention^13^. The use of TXA in the current study was linked to a significant decrease in the total volume of postoperative drainage and a reduced period of drainage. The use of TXA was also linked to a much lower incidence of hematoma and numerically lower rates of seroma and surgical-site infection, but the latter differences were not statistically significant.

Past research has shown that TXA has the potential to decrease the complication of hematoma in breast surgery, such as reduction mammoplasty and implant-based reconstruction^14–17^. In line with our results of lesser perioperative reductions in Hb and Hct (Table 3), the current results are in line with the earlier studies by Weissler et al., who found a reduced hematoma occurrence after the use of TXA in implant-based breast reconstruction^11^. It is important to note that no patients in the TXA group had a hematoma, and six patients in the control group had to be operated on because of hematoma. We hypothesize that the more postoperative drainage in the first 24 hours, the higher the risk of subsequent hematoma formation, which could be a manifestation of more perioperative bleeding. Nevertheless, this hypothesis needs to be validated in bigger prospective studies.

The risk of postoperative seroma can be minimized by careful flap dissection with minimal tissue trauma and less operative time^18,19^. We have implemented two measures to reduce the risk of seroma after breast cancer surgery based on our surgical experience. First, it is possible that preservation of the adipose tissue under the extended ELD muscle during flap harvest can preserve the integrity of the local lymphatic network and, thus, decrease the postoperative accumulation of fluid^20^. Second, intravenous TXA can decrease intraoperative bleeding and possibly decrease the chances of seroma development by enabling more effective hemostasis and, therefore, reducing the duration of the operation^21^. The difference was not statistically significant, but patients receiving TXA showed a tendency towards a lower incidence of seroma compared to the control group. Notably, conservative management, such as immobilization, was effective in all patients who developed seromas, and they healed within 1 month without further surgical intervention.

Long drainage and continued postoperative fluid build-up can predispose to bacterial colonization and surgical-site infection. In the current research, intravenous TXA was linked to reduced postoperative drainage volumes and reduced period of drain placement. Lohani et al. found a reduced rate of wound-related infections in patients treated with TXA, which was partly due to the earlier removal of drains and the decreased development of seromas^20^. We have found that our results are in line with other studies that have shown that TXA has the ability to decrease the duration of postoperative drainage and the overall drainage volume. Moreover, the fact that the percentage of patients whose drains were taken out in 10 days was higher in the TXA group (Table 2, Panel A) is another evidence of this observation.

TXA had a significant effect on the drain output in the first 24 hours of the postoperative period but not in the next 5 days. This finding is consistent with previous reports by Sahinovic, who similarly observed that the drain-reducing effect of TXA was most apparent during the early postoperative period^22^. This time-dependent trend indicates that the hemostatic effect of TXA might be the strongest in the early postoperative period when bleeding and exudative fluid buildup are the highest. Thus, use of TXA can offer specific short-term advantages in the acute phases of wound healing by decreasing the initial fluid build-up and enabling the removal of the drain earlier.

Prudovsky et al. reported that TXA can regulate inflammatory responses by its antifibrinolytic activity, and systemic inflammatory responses have been reduced in experimental animal models^23^. The current research showed that the postoperative CRP levels were significantly lower in the TXA group, which may indicate a potentially suppressed systemic inflammatory response. In addition to its canonical action in hemostasis, TXA also competitively binds the lysine-binding sites of plasminogen, thus preventing its conversion to active plasmin^24^. Plasmin may mediate inflammatory cross-talk by cleaving complement proteins, such as C3 and C5, and activating immune cells, such as monocytes and macrophages, to secrete pro-inflammatory cytokines, such as interleukin-6 (IL-6)^25–27^. Thus, the plasmin activity inhibition by TXA could suppress these inflammatory pathways and, as a result, the synthesis of acute-phase reactants like CRP. Because CRP was the only measured inflammatory biomarker in the current study, more prospective research including several inflammatory biomarkers is needed to differentiate between direct immunomodulatory action of TXA and indirect action of decreased surgical trauma and hematoma development.

No significant rise in thrombotic complications after the use of TXA in free-flap or implant-based breast reconstruction has been reported in previous studies. In line with these results, there were no symptomatic DVT events in either group in the current study. Nevertheless, due to the absence of DVT events and postoperative enoxaparin prophylaxis in all patients, our study could not identify whether the use of TXA affects thromboembolic risk. Thus, the lack of thromboembolic events cannot be viewed as a conclusive sign of the thrombotic safety of TXA in this context.

A major limitation of the current study is the possibility of temporal confounding. Since TXA was regularly implemented at one calendar time point (July 2021), the exposure to TXA was intrinsically linked to time. Although multivariate analyses were conducted to control for age, BMI, type of reconstruction, axillary procedure, and neoadjuvant chemotherapy, the covariates are not able to completely separate the effects of TXA and secular changes in perioperative care, surgical practice, or other unmeasured variables. Therefore, the associations observed can be viewed as possibly affected by both TXA exposure and time trends.

There are a number of other limitations that must be recognized. First, the retrospective, single-center design and rather small sample size restrict the extrapolation of the results and make it impossible to draw definite causal conclusions. Second, retrospective data collection could have led to incomplete reporting of seroma events, especially small or clinically insignificant collections that were treated conservatively outside of the hospital. Third, due to the fact that all patients were given postoperative enoxaparin prophylaxis and no symptomatic DVT events were observed, the study lacked the power to assess the relationship between TXA administration and thromboembolic risk. Lastly, CRP was the sole inflammatory biomarker assessed, which restricted the interpretation of the possible immunomodulatory effects of TXA. Future multicenter, ideally randomized, standardized follow-up, systematic evaluation of postoperative complications, extensive inflammatory profiling, and proper surveillance of thromboembolic events are justified to confirm the effectiveness and safety of TXA in immediate ELD flap breast reconstruction.

## Conclusion

In conclusion, intravenous administration of TXA was linked to decreased postoperative drainage volume, decreased drainage time, and a decreased rate of clinically significant hematoma in immediate ELD flap breast reconstruction. Though these results indicate that TXA can have significant perioperative advantages in this context, the retrospective, single-center nature and the possibility of temporal confounding restrict causal inference. Thus, the current results are to be viewed as hypothesis-generating, but not as conclusive evidence of a causal treatment effect. More prospective, multicenter randomized controlled trials are justified to confirm these findings and determine the effectiveness and safety of TXA in immediate ELD flap breast reconstruction.

## Data Availability

Aggregate results are reported in the manuscript. The individual-level patient data underlying this study are not publicly available due to patient confidentiality.

